# Artificial Intelligence for Anatomical Structure Identification on Ultrasound in Regional Anaesthesia: A Scoping Review Protocol

**DOI:** 10.1101/2023.07.04.23291560

**Authors:** James S Bowness, David Metcalfe, Kariem El-Boghdadly, Neal Thurley, J Alison Noble, Helen Higham

**Author notes:** **Corresponding Author** James S Bowness, OxSTaR Centre, Nuffield Division of Anaesthetics, Nuffield Department of Clinical, Neurosciences, John Radcliffe Hospital, Oxford, OX3 9DU.

## Abstract

**Background:** Ultrasound is the most common form of guidance for regional anaesthesia. There is increasing interest in developing supporting technology, particularly in the form of artificial intelligence (AI), to aid in the acquisition and interpretation of optimal ultrasound views for these procedures. However, this is a broad field, with academia, clinical practice, and industry all providing disparate contributions. We will undertake a scoping review of publicly-available data, to assess methods of evaluation for accuracy and utility of such systems.

**Methods:** We will perform searches in multiple databases, including ACM Digital Library, CINAHL, EMBASE, IEEE Explore, and OVID MEDLINE. We will search the International Committee of Medical Journal Editors approved clinical trial registries and the World Health Organisation (WHO) clinical trials registry for studies registered in this field. Grey literature will be searched through the online library of doctoral theses (http://ethos.bl.uk/Home.do), regulatory authority registries and competent authority websites of North America and the UK, the websites of international learned societies in regional anaesthesia, and material from commercial organisations with products in the field.

The primary goal is to summarise the approaches used to evaluate accuracy and utility of these devices. A secondary goal is to assess the standardisation of reporting in this field, with particular reference to whether reporting guidelines have been followed.

**Discussion:** To the best of our knowledge, this is the first scoping review of this type. Synthesis of the available evidence will enable us to make recommendations on standardised evaluation approaches of assessment, to allow robust and relevant evaluation which can be compared to similar evaluations of other devices.

## Background

Ultrasound image guidance for regional anaesthesia was first described in 1989.^1^ It has since become the main method to guide needle insertion and direct the delivery of local anaesthetic to inhibit the action of specific peripheral nerves.^2, 3^ Ultrasound-guided regional anaesthesia (UGRA) intends to use ultrasound to identify relevant sono-anatomy, to aid in directing the needle toward the desired target(s) and avoid unwanted trauma of important proximate structure(s).^4-6^ It is also utilised to image the spine and surrounding tissues ahead of performing central neuraxial blockade (CNB; spinal and epidural).^7^ Fundamental to this is a thorough knowledge of the underlying anatomy, which informs the appropriate acquisition and interpretation of ultrasound images.^6, 8, 9^ However, anaesthetists’ knowledge of clinical anatomy is variable,^10^ ultrasound is known to be an operator-dependent imaging modality,^9^ and fallibility in human interpretation of medical images is recognised.^11^

Artificial intelligence (AI) is becoming a mainstream methodology for medical image interpretation. It is a general term which refers to the ability of computers to perform tasks traditionally associated with human intelligence.^12^ Computer vision – a field dealing with the ability of computers to interact with the visual world – often involves the use of machine learning (ML). This includes techniques which a enable computer programs to improve performance for a given task (learn) with increased exposure to training data. Deep learning (DL), a field within ML, uses artificial neural networks (interconnected mathematical functions) arranged in layers, allowing progressively more information to be extracted from the input data at each layer. Examples of AI-based ultrasound image analyses are emerging in clinical practice.^13,14^

Deep learning for image analysis involves developing an algorithm (a rule-based problem-solving function executed by a computer) and exposing it to training data. These training data can be ‘labelled’ with the correct answer or features of interest (supervised ML) or presented alone (unsupervised ML). In supervised ML, as more training data is presented to the algorithm, the modifiable (learnable) parameters of the algorithm change as the algorithm ‘learns’ to make associations between the input data and training labels. When training is completed, and the fixed algorithm is deployed in its desired use case, it can be used to predict the appropriate label(s) for new image data. In unsupervised ML, the training data are presented without paired labels. The algorithm learns to identify clusters in the data (areas of similarity) that can then be assigned to a category (or label). Training stops when the identification of these clusters is considered suitable for the desired use case, and the algorithm can be deployed in the same manner as with supervised ML.

Recent publications present the case for the use of AI for ultrasound scanning in UGRA; peripheral nerve blocks (PNB) and CNB.^15, 16^ Potential benefits may include aiding the performance of ultrasound scanning in regional anaesthesia, training and learning these skills, and facilitating the use of UGRA outside the traditional environment of anaesthesia (e.g., the emergency department).^15, 17^ There are few systems with regulatory approval for clinical use and this is an emerging field – the first system gained regulatory approval for clinical use in PNBs in April 2021.^18^ Recently, academic reporting guidelines for studies involving medical AI devices have been published,^19, 20^ but these make little recommendation as to how studies should be structured. Also, there are no formal standards which guide consistency in regulatory approval of these devices. Thus, evaluations of such devices for the identification of anatomical structures on ultrasound in relation to UGRA are heterogeneous. Available data include proof-of-concept studies in pre-clinical, experimental work^16, 21, 22^ to early assessments of commercially-available systems.^17, 23-25^

Furthermore, this is an interdisciplinary field with expertise drawn from computer science, engineering, and clinical medicine. Specialists may focus on different endpoints or use different approaches to assess the same endpoint. As such, opportunities for communication between clinicians, anatomists, computer scientists, engineers, mathematicians, and industry/manufacturers are likely to be missed. There is no single source of information which brings together different thought processes in this field to analyse early studies and assess areas of strength or weakness. Importantly, there are few recommendations on how this field can move forward, with a particular focus on the consistent and standardised evaluation of such new devices.

Thus, it is important to survey the literature and combine the findings into a coherent summary. This scoping review aims to identify and map the available evidence in this field. It is intended to inform future work by identifying areas where development is needed and proposing a formal, standard assessment framework which future evaluation processes can follow. Data included from adult subjects and from the point of introduction of ultrasound image guidance in regional anaesthesia (1989) will be considered.

A preliminary search (conducted on 4^th^ May 2022) was unable to identify scoping or systematic reviews registered with the Joanna Briggs Institute (JBI) Database of Systematic Reviews and Implementation Reports (https://csu-sfsu.primo.exlibrisgroup.com/discovery/fulldisplay?vid=01CALS_SFR:01CALS_SFR&tab=jsearch_slot&docid=alma991067665362702901&searchScope=EVERYTHING&context=L&lang=en), Cochrane Database of Systematic Reviews (https://www.cochranelibrary.com/cdsr/reviews), or the International Prospective Register of Systematic Reviews (PROSPERO; https://www.crd.york.ac.uk/prospero).

### Scoping Review Objectives

The objective of this review is to provide an overview of the available evidence on the accuracy and utility of artificial intelligence (AI) systems for ultrasound scanning in regional anaesthesia. This will be used to highlight areas of strength and deficiency in the current body of knowledge, and propose the concept of a formal, standardised assessment framework which future evaluation processes can follow. This will provide a basis for further research using similar technologies in this nascent and rapidly developing field.

## Methods

### Protocol and Registration

The protocol presented below was developed using the guidance provided by the JBI and Preferred Reporting Items for Systematic Reviews and Meta-Analyses – Extension for Scoping Reviews (PRISMA-ScR).^26-28^

The review was registered with the JBI Database of Systematic Reviews and Implementation Reports (https://csu-sfsu.primo.exlibrisgroup.com/discovery/fulldisplay?vid=01CALS_SFR:01CALS_SFR&tab=jsearch_slot&docid=alma991067665362702901&searchScope=EVERYTHING&context=L&lang=en) prior to data extraction from the identified sources.

### Scoping Review Question

“What is the evidence supporting the use of artificial intelligence for ultrasound scanning in regional anaesthesia?”

### Inclusion Criteria

#### Population

Sources must be published during/after 1989 – the date of the first publication relating to ultrasound image guidance in regional anaesthesia.^**1**^ Sources must pertain to data gathered from human ultrasound scanning (living or cadaveric studies). Data from animal and bench studies will be excluded.

#### Concept

Data assessed will relate to the accuracy of AI-based anatomical structure identification on real-time B-mode ultrasound and the utility of these systems with respect to clinical practice. For the purposes of this review, utility refers to data related to anaesthetists’ ultrasound scanning performance and patient outcomes (efficacy and safety of UGRA).

#### Context

Sources must relate to ultrasound scanning in the context of CNB (e.g., spinal and epidural) and PNBs.

#### Types of sources

All published research (including academic publications, clinical/specialist society information, and commercial product literature) will be considered. Sources published in languages other than English will be translated and/or the author(s) will be contacted for assistance with data extraction.

### Search Strategy

#### Published Academic Literature

This search strategy was developed by the lead investigator (JSB) and reviewed/modified by the team (including a medical librarian, NT, who will execute the academic searches).

It will utilise the ACM Digital Library, CINAHL, EMBASE, IEEE Explore, and OVID MEDLINE. Searches will be limited by date (1989 – present) and subjects scanned (human only, exclude animal studies).

An initial search of two databases (OVID Medline and EMBASE) will be performed as shown in Table 1. Screening of manuscript titles and abstracts will be performed (as described below). The key words of included manuscripts will be reviewed and added to the search strategy, if not already included.

**Table 1.** Initial search strategy.

| Artificial Intelligence | AND | Ultrasound | AND | Regional Anaesthesia |
| --- | --- | --- | --- | --- |
| “artificial intelligence”.ti,ab. |  | ultraso*.ti,ab. |  | anaesthe*.ti,ab. |
| OR |  | OR |  | OR |
| AI.ti,ab. |  | sonog*.ti.ab |  | anesthe*.ti,ab. |
| OR |  |  |  | OR |
| “computer vision”.ti,ab. |  |  |  | regional.ti,ab. |
| OR |  |  |  | OR |
| “machine intelligence”.ti,ab. |  |  |  | nerve*.ti,ab. |
| OR |  |  |  | OR |
| “deep learning”.ti,ab. |  |  |  | block*.ti,ab. |
| OR |  |  |  | OR |
| “machine learning”.ti,ab. |  |  |  | spinal.ti,ab. |
| OR |  |  |  | OR |
| “neural network*”.ti,ab. |  |  |  | epidural.ti,ab. |
| OR |  |  |  | OR |
| “computer aid*”.ti,ab. |  |  |  | “central neuraxial”*.ti,ab. |
| OR |  |  |  |  |
| automat*.ti,ab. |  |  |  |  |
| OR |  |  |  |  |
| segment*.ti,ab. |  |  |  |  |

The full search will then be executed on all databases. The next stage of the search strategy will involve scrutiny of all relevant cited and citing work of the publications included after screening.

#### Other Literature

As AI is an area of intense commercial research, other evidence and commercial data will also be sought to minimise publication bias. Due to the variable search capability of these sources, the search terms for these repositories will use the subject headings (and all combinations thereof) from the search strategy of this scoping review to enable a consistent search approach (“artificial intelligence; ultrasound; anaesthesia [anesthesia]”).

The International Committee of Medical Journal Editors (ICMJE) approved clinical trial registries will be searched (www.anzctr.org.au; www.clinicaltrials.gov; www.ISRCTN.org; www.umin.ac.jp.ctr; www.trialregister.nl; https://eudract.ema.europa.eu/). As the protocols and data from EudraCT are available via the EU Clinical Trials Register (www.clinicaltrialsregister.eu/), this will be searched in its place. In addition, the World Health Organisation (WHO) clinical trials registry platform (http://www.who.int/ictrp/search/en/) will be searched.

The online library of doctoral theses (http://ethos.bl.uk/Home.do) will also be searched, as will regulatory authority registries and competent authority websites of North America (US Food & Drug Administration; MAUDE, 510k, De Novo, and Medical Device Recall databases) and the UK (Medicines and Healthcare products Regulatory Agency).

To review centralised information in the clinical specialty, two investigators will review websites of prominent, international learned societies in regional anaesthesia for material relating to AI in UGRA:

- African Society for Regional Anesthesia (AFSRA; http://afsra.org)
- American Society of Regional Anesthesia & Pain Medicine (ASRA; https://www.asra.com)
- Asian and Oceanic Society of Regional Anaesthesia and Pain Medicine (AOSRA; https://aosrapm.org)
- European Society of Regional Anaesthesia and Pain Therapy (ESRA; https://esraeurope.org)
- Latin American Society of Regional Anesthesia (LASRA; www.lasra.com.br)
- Regional Anaesthesia UK (RA-UK; https://www.ra-uk.org)

Material from commercial organisations with products in the field will also be reviewed, including Intelligent Ultrasound (https://www.intelligentultrasound.com), Smart Alpha (https://www.nerveblox.com), GE Healthcare (https://www.gehealthcare.co.uk), Mindray (https://www.mindray.com), and Samsung (https://www.samsung.com).

### Data Extraction

#### Screening Initial Results

Once all sources have been identified from this search, two investigators will screen the titles for inclusion in the study. Duplicates will be removed, and the titles will then be assessed to determine whether they relate to AI, ultrasound and regional anaesthesia. If the title is ambiguous, the source will be included in the abstract review. In case of discrepancy between the investigators’ assessments, a third investigator will review the title and adjudicate.

#### Abstract Review

A reviewer will the review the abstract/summary of the included sources to consider inclusion for a full-text review. Sources will be included if the abstract/summary relates to AI, ultrasound, and regional anaesthesia (on human subjects).

#### Extraction of Results

The full text of all appropriate sources will be reviewed to ensure that each is still deemed suitable. Results and conclusions will be scrutinised to extract data on accuracy and/or utility of the AI systems evaluated. The extracted data will be electronically charted using Table 2.

**Table 2.** Data Extraction Form.

|  |
| --- |
| Study ID |
| Details of Publication |
| Title |
| Author(s) |
| Year |
| Country of origin |
| Language of source |
| Type of source:<br>• Academic (full paper/abstract/unpublished and field of publication/authors e.g., computer science, clinical)<br>• Other literature (e.g., trials registry, industry material) |
| Nature of Work |
| Reported according to formal guidelines? (E.g., CONSORT-AI, DECIDE-AI) |
| Aim of Assessment (accuracy or utility) |
| New technology or validate findings from earlier study(ies)? |
| Intervention<br>• AI methodology (techniques used)<br>• Task of AI system (e.g., bounding box, segmentation, tracking)<br>• Training data (e.g., type/size of training dataset, including scan subjects/images & who labels data)<br>• Comparison (e.g., to human expert) |
| Population Assessed:<br>• Number of subjects<br>• Subject demographics (age, gender, BMI) |
| Type/size of test dataset (images/videos) |
| Outcomes:<br>• Accuracy - clinical (e.g., expert opinion) or non-clinical (e.g., Dice metric)<br>• Utility |
| Key Findings that relate to this Scoping Review |
| Any Additional Information |

This is a scoping review of data in an emerging field. Therefore, the investigators have not predefined what data may pertain to accuracy or utility, or set any minimum outcome/reporting criteria. To maximise data capture, an inclusive approach will be adopted when reviewing publications. For example, utility may include UGRA efficacy, safety, cost-effectiveness, implementation etc. As sources of commercial data may not include methodological reporting, this is not a prerequisite for inclusion. The sources may refer to ultrasound scanning, in a simulation or clinical setting, as the scanning itself is performed on human tissue.

### Collating and Summarising Data, Presentation of Results

The search and screening process will be shown in a flowchart that details the number of publications considered at each stage.

The data extracted from this search and review process will be presented in a descriptive format in two parts. One element will include a discussion of data pertaining to the accuracy of AI systems in identifying anatomical structures on ultrasound. The investigators anticipate differing forms of data and analyses in this element; thus, these forms will be summarised and the range of accuracy(ies)/methods will be presented. The second element will discuss data pertaining to clinical utility: improving the performance of anaesthetists’ ultrasound scanning (e.g., the anaesthetist’s ability to identify structures) or improving patient outcomes.

## Discussion

The proposed scoping review aims to summarise the available evidence on the accuracy and utility of AI systems for ultrasound scanning in regional anaesthesia.

The amount and content of available data are likely to reflect that this is an emerging field, and this study may highlight a lack of consistency between studies. Therefore, we hope this review can serve as a concise summary of the state of the art and provide guidance for the direction and structure of future works.

## Limitations

Despite the systematic search strategy summarised above, we anticipate challenges in the proposed study. We recognise the possibility of publication bias in the available work and that studies conducted as part of theses may not be found, despite the thorough search strategy. Given the pace of progress in this sector, further studies may be in progress but not in the public domain. Industry data may not be available, intentionally or otherwise, as companies may not always have a commercial incentive to share data.

As there are few systems available in clinical practice, and the first of these was only approved in 2021,^29^ we anticipate that a large number of the included sources will be proof-of-concept studies with few scan images/subjects. It is likely that few, if any, will include patient outcome data.

## Data Availability

No data is available in this manuscript (review protocol only)

## Notes

**Conflict of Interest Statement** JSB is a Senior Clinical Advisor for Intelligent Ultrasound, receiving research funding and honoraria. KEB is an Editor for *Anaesthesia* and has received research, honoraria and educational funding from Fisher and Paykel Healthcare Lit, GE Healthcare and Ambu. JAN is a Senior Scientific Advisor for Intelligent Ultrasound.

### Competing Interest Statement

JSB is a Senior Clinical Advisor for Intelligent Ultrasound, receiving research funding and honoraria. JAN is a Senior Scientific Advisor for Intelligent Ultrasound.

### Funding Statement

This study did not receive any funding

